# Fully Quantitative Measurements of Differential Antibody Binding to a Spectrum of SARS-CoV-2 Spike Proteins: Wuhan, Alpha, Beta, Gamma, Delta, Omicron BA.1, BA.4, BA.5, BA.2.75 and BA.2.12.1

**DOI:** 10.1101/2023.01.11.23284431

**Authors:** Philip H. James-Pemberton, Shivali Kohli, Aaron C. Westlake, Alex Antill, Jade Hunt, Rouslan V. Olkhov, Andrew M. Shaw

## Abstract

A fully quantitative comparative analysis has been performed on the differential antibody binding to a spectrum of Spike proteins to the SARS-CoV-2 variants Wuhan, Alpha, Beta, Gamma, Delta and Omicron BA.1, BA.1, BA.4, BA.5, BA.2.75 and BA.2.12.1. The immunity profile was determined for four patient cohorts: pre-pandemic, the first infection in the pandemic, Wuhan(+), and two vaccinated cohorts, the initial double-vaccination with AstraZeneca (AZ) and Pfizer and a final boosted cohort including with known vaccination but unknown mixture of natural infection. A universal protection immunity endotype, U(+), with significant antibody levels to all ten variants was observed in with a incidence of 11% (95% CI 4% - 25%) in the Wuhan(+) cohort challenging directly the ‘one-and-done’ immunity claim. The U(+) incidence rises to 22% (95% CI 12% - 37%) in the double-vaccinated cohort and 54% (95% CI 39% - 68%) in the triple vaccinated cohort. The remaining patients in each cohort show a spectrum of immunity with some drop-out immunity endotypes, U(±), showing poor antibody response to one or more variants. The U(±) incidence in the triple vaccination cohort is 41% (95% CI 28% - 57%) suggesting patients with poor sterilising sera may not clear a SARS-CoV-2 infection leading to viral persistence and mobile microcolonies that may provide a pathophysiology for the symptoms of long Covid.

**Funding:** Exeter University Alumni, Attomarker Ltd-funded PhD studentship (PJP) at the University of Exeter and Attomarker Ltd funding directly.

## Introduction

The prevalent variants of SARS-CoV-2 in 2022 have been Omicron sub-variants leading to the use of bi-valent mRNA vaccines to produce spike proteins for both Wuhan and the Omicron sub-variants, BA.5^1^ (Pfizer) and BA.1^2^ (Moderna). In a previous study^3^ we investigated the antibody response profile to spike proteins from the SARS-CoV-2 variants Wuhan, Alpha, Beta, Gamma, Delta and Omicron BA.1. This study extends the spike protein spectrum to include a set of Omicron variants BA.4, BA.5, BA.2.75 and BA.2.12.1 to give a ten-dimensional antibody response to variant spike proteins (Figure S1). Mutations to the receptor binding domain (RBD) region of the spike protein are reported to improve binding to the ACE2 receptor; the binding event necessary for infection and thus enhanced infection rates^4^. Further, mutations to the RBD have reduced the effectiveness of neutralising antibodies targeted at this region indicating reduced vaccine efficacy^5-7^. Further, the BA.1 – BA.5 mutations show increased potential for vaccine escape^8^ and the consequent loss of effectiveness of monoclonal antibody treatments ^9^. However, some studies using different vaccine combinations^10,11^ have shown some improvement in the resulting neutralising antibody protection, and the new bivalent vaccines can provide better neutralising antibody binding^2^.

Previously, we identified a number of immunity endotypes based on antibody binding to the whole spike protein, including the RBD region, and identified a universal endotype, U(+), with antibodies to a conserved epitope across the variants. We identified a possible hinge region on the spike protein that is critical to the function at docking and a transition from the pre-fusion to fusion state upon binding to the ACE2 receptor. Antibodies to this ‘Achilles epitope’ would remain active across the variants as mutation to the hinge could result in complete loss of function.

In this paper, we determine the fully quantitative endotype analysis for a spectrum of SARS-CoV-2 variants: Wuhan, Alpha, Beta, Gamma, Delta, Omicron BA.1, BA.4, BA.5, BA.2.75 and BA.2.12.1 using multiplexed assays. Standardisation of immunoassay measures is possible for biophotonic sensor platforms based on the NIST standard human antibody^12^ specific to the respiratory syncytial virus protein F (RSVF). First, the different variant proteins are screened with a panel of antibodies to establish epitope integrity for the calibration site in the S2 region of the protein as well as epitope degradation in the S1 region. Patient samples were then screened from the same four cohorts: from pre-pandemic (controls); from the Wuhan wave pre-vaccine; and fully vaccinated and boosted samples for select combinations of vaccination. The endotype analysis is extended over all ten variants to assess conservation of the epitope for the Universal endotype U(+) and the incidence of Omicron drop-out immunity endotypes.

## Methods and Materials

### Methods

#### Biophotonic Multiplexed Immuno-kinetic assay

A biophotonic platform using localised particle plasmon resonance has been described in detail elsewhere in a SARS-CoV-2 antibody sensing application^13^. The resulting assays are fully quantitative and calibrated against the NIST standard human antibody, RM8671, NISTmAb, a recombinant humanized IgG1? with a known sequence^14^. The spike calibration antibody (40590-D001, Sinobiological) is targeted at a conserved region of the S2 region of the protein to assess the integrity of all variant proteins.

The endotype classification was performed manually using a simple selection hierarchy algorithm leading to definitions, Table S1. The classification is based on the instrument characteristics: Limit of Detection (LoD) = 0.2 mg/L and 10*×*LoD = Limit of Quantification (LoQ) = 2 mg/L. A full correlation analysis for all response combinations was also performed, from which the correlation coefficients and lines of best fit were derived.

The incidence in each of the endotype groups is calculated as a simple percentage proportion with Wilson 95% confidence limit estimates based on sample size. Significant of median difference between the cohorts was assessed using the Wilcoxon test.

### Materials

Materials used throughout the course of the experiments were used as supplied by the manufacturer, without further purification. Sigma-Aldrich supplied phosphate buffered saline (PBS) in tablet form (Sigma, P4417), phosphoric acid solution (85 ± 1 wt. % in water, Sigma, 345245) and Tween 20 (Sigma, P1379). Glycine (analytical grade, G/0800/48) was provided by Fisher Scientific. Assay running and dilution buffer was PBS with 0.005 v/v % Tween 20 and the regeneration buffer was 0.1 M phosphoric acid with 0.02 M glycine solution in deionized water.

The recombinant Human Antibody to the Spike protein S2 subdomain was a chimeric monoclonal antibody (SinoBiological, 40590-D001, Lot HA14AP2901). The antibody was raised against the following immunogen: recombinant SARS-CoV-2 / 2019-nCoV Spike S2 ECD protein (SinoBiological, 40590-V08B).

NISTmAb, Humanized IgG1κ Monoclonal Antibody from National Institute of Standards and Technology (RM8671). The NISTmAb is a recombinant humanized IgG1? with a known sequence^14^ specific to the respiratory syncytial virus protein F (RSVF)^15^. The detection mixture consisted of a 200-fold dilution of IG8044 R2 from Randox in assay running buffer.

Two sensor chips designs were printed; an Omicron focussed array with recombinant human serum albumin (rHSA) from Sigma-Aldrich (A9731), protein A/G (PAG) from ThermoFisher (21186), and six SARS-CoV-2 Spike Protein variants for the Wuhan and trimer proteins for Omicron BA.1, BA.2.12.1, BA.4, BA.5 strains from SinoBiological and BA.2.75 spike protein from Acro Biosystems as detailed in Table S2. The other sensor chip design included earlier variants, with rHSA, PAG and six monomer Spike Protein variants: Wuhan, Alpha, Beta, Gamma, Delta and Omicron BA.1, also detailed in Table S2.

### Patient Samples

#### Commercial samples

Serum samples were purchased from two suppliers (Biomex GmbH and AbBaltis). 17 pre-pandemic (pre-December 2019), PCR(-) human serum samples were purchased from AbBaltis. All were tested and found negative for STS, HbsAg, HIV1 Ag (or HIV PCR(NAT)), HIV1/2 antibody, HCV antibody and HCV PCR(NAT) by FDA approved tests. 11 positive samples, all from PCR(+) individuals, were purchased from AbBaltis.. No information was provided regarding symptoms of the donors. 45% of these samples were from female donors and 55% were from male donors. The age of donors ranged from 19 to 81 years. No information on time from infection to sample collection was given.

Samples purchased from Biomex (*n*=26) were from PCR(+) individuals. All samples were YHLO Biotech SARS-CoV-2 IgG positive and Abbott SARS-CoV-2 IgG positive. A spectrum of patient symptoms from the following list were detailed for each sample: fever, limb pain, muscle pain, headache, shivers, catarrh, anosmia, ache when swallowing, diarrhoea, breathing difficulties, coughing, tiredness, sinusitis, pneumonia, sickness, lymph node swelling, pressure on chest, flu-like symptoms, blood circulation problems, sweating, dizziness, and hospitalisation. Time from infection to sample collection ranged from 27 to 91 days. 35% of these samples were from female donors and 65% were from male donors. The samples were collected prior to June 2020, so PCR(+) samples will result from infection by the SARS-CoV-2 strains circulating prior to this time, most similar to the Wuhan protein. These commercial samples, 36(+) and 16(-), were tested using the Spike Variant Array as a part of this study.

#### Attomarker Clinic samples

Samples were collected from patients in Attomarker clinics, all of whom provided informed consent for their anonymised data to be used in research to aid the pandemic response. The data from tests of 96 patient samples are included in this study: 13 had received one dose of either AstraZeneca (AZ) or Pfizer SARS-CoV-2 vaccine; 41 had received two doses of either AZ, Pfizer or Moderna SARS-CoV-2 vaccine at least 14 days prior to sample collection and testing by Attomarker; 41 further samples were from patients who had received a third vaccination. Full patient demographics are shown in Table *1*. One sample had no vaccine history disclosed.

**Table 1.**
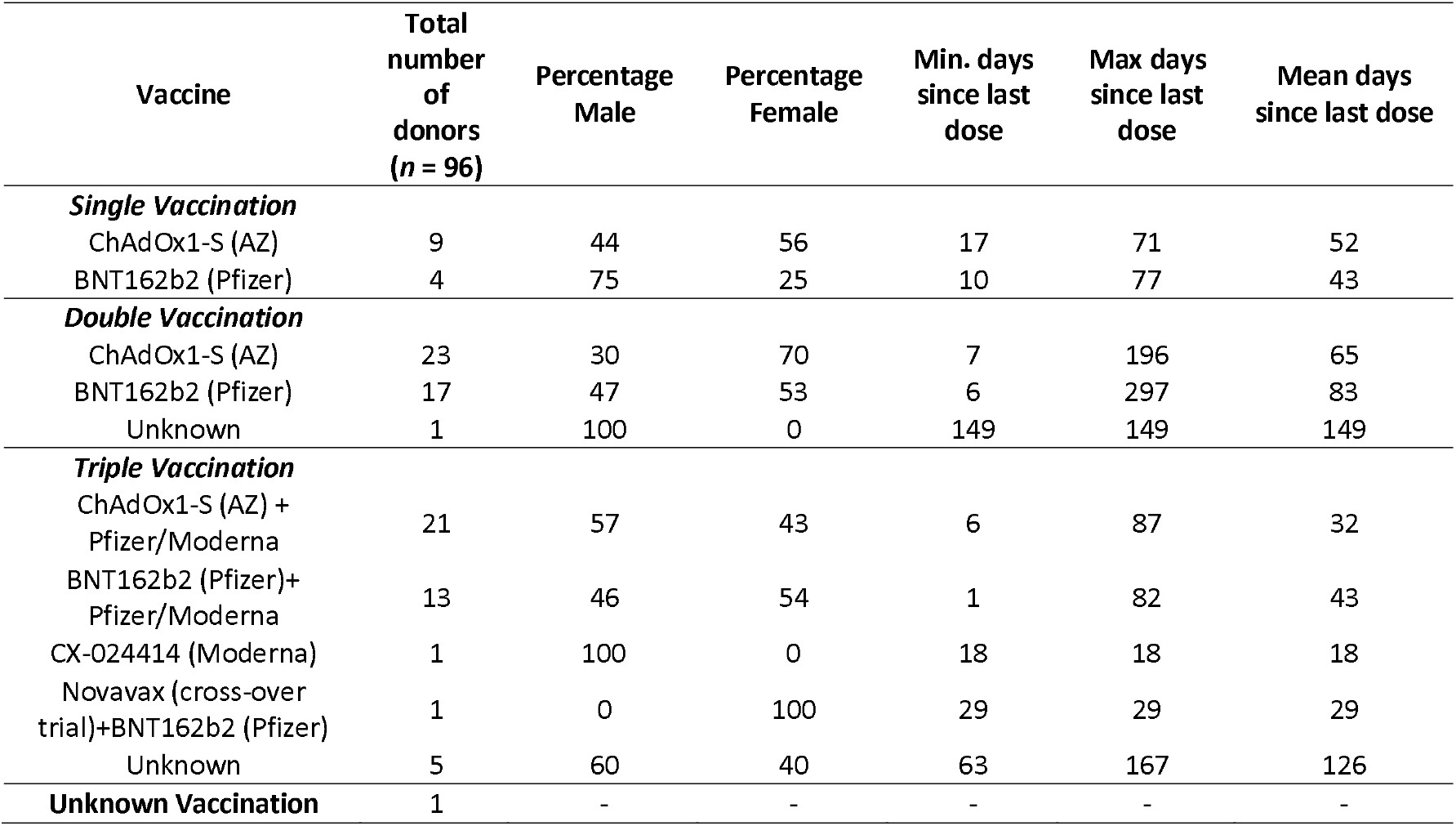
Demographic data for samples from vaccinated individuals collected in the Attomarker clinic still not right!

| Vaccine | Total number of donors ($n = 96$) | Percentage Male | Percentage Female | Min. days since last dose | Max days since last dose | Mean days since last dose |
| --- | --- | --- | --- | --- | --- | --- |
| **Single Vaccination** |  |  |  |  |  |  |
| ChAdOx1-S (AZ) | 9 | 44 | 56 | 17 | 71 | 52 |
| BNT162b2 (Pfizer) | 4 | 75 | 25 | 10 | 77 | 43 |
| **Double Vaccination** |  |  |  |  |  |  |
| ChAdOx1-S (AZ) | 23 | 30 | 70 | 7 | 196 | 65 |
| BNT162b2 (Pfizer) | 17 | 47 | 53 | 6 | 297 | 83 |
| Unknown | 1 | 100 | 0 | 149 | 149 | 149 |
| **Triple Vaccination** |  |  |  |  |  |  |
| ChAdOx1-S (AZ) + Pfizer/Moderna | 21 | 57 | 43 | 6 | 87 | 32 |
| BNT162b2 (Pfizer)+ Pfizer/Moderna | 13 | 46 | 54 | 1 | 82 | 43 |
| CX-024414 (Moderna) | 1 | 100 | 0 | 18 | 18 | 18 |
| Novavax (cross-over trial)+BNT162b2 (Pfizer) | 1 | 0 | 100 | 29 | 29 | 29 |
| Unknown | 5 | 60 | 40 | 63 | 167 | 126 |
| **Unknown Vaccination** | 1 | - | - | - | - | - |

#### Ethical Approval

The use of the Attomarker clinical samples was approved by the Bioscience Research Ethics Committee, University of Exeter.

## Results

The combined patient cohorts showed a range of immunity endotypes in response to the spectrum of SARS-CoV-2 spike proteins. The pre-pandemic, negative control samples were 81% spike antibody negative or unclassified with 2 patients showing a *β*/*γ*/*δ* dropout suggesting immunity to Wuhan and the Omicron sub-variants potentially due to a misclassification: the PCR false-positive rate was typically 4%. One patient (6% (95% CI 1% - 28%)) had a α/γ/δ/2.75/4/5/2.12.1/W dropout but was positive for Beta and Omicron BA.1: this may result from alphacoronavirus or random antibody cross-reactivity.

However, for the W(+), double vaccinated and triple-vaccinated cohorts there was a significantly larger spectrum in antibody responses, Figure 1, with the full classification shown in Table 3. The ideal antibody response to a universal epitope present in all variant spike proteins, U(+) Figure 1A, was observed in 11% (95% CI 4% - 25%) of the immunologically naïve Wuhan(+) patients, whilst 53% (95% CI 37% - 68%) of patients presented with one or more dropouts, U(±). The most prevalent U(±) was a triple drop-out, Figure 1D, *β*/*γ*/*δ*(-). The U(+) incidence increases for the double-vaccinated cohort 22% (95% CI 12% - 37%) and rises again in the triple-vaccinated cohort to 54% (95% CI 39% - 68%). The double-vaccine groups for AZ or Pfizer offer different protection across the spectrum of variants with AZ showing U((1-8)-) dropouts dominated by U(5-) including the Omicron sub-variants, Figure 1 (F,G,H,I). The Pfizer vaccine by comparison only has U(1-4) including a full mix of all variants. In the triple-vaccination cohort the higher-order drop-outs are nearly eliminated with no U(5) or greater. One patient with a triple vaccination also had an octuple drop-out, Figure 1I, suggesting a lack of immunity across all variants.

**Figure 1.**
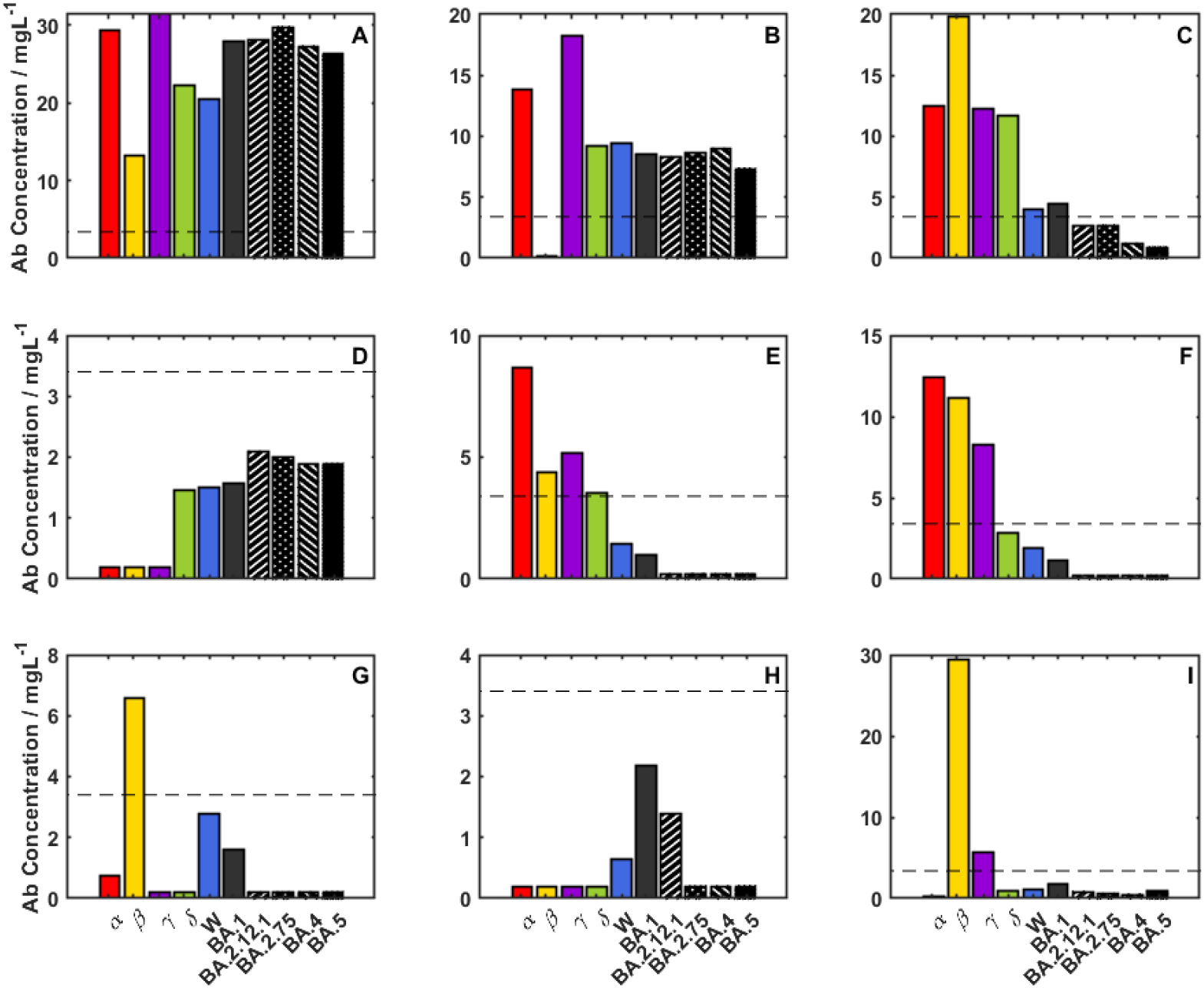
Immunity Endotype examples from the Omicron variants: (A) a universal positive, U(+) [2× Pfizer], (B)Single Dropout β(-) [2× Pfizer], (C) Double Dropout [W(+)], (D), Triple Dropout [W(+)], (E) Quadruple Dropout [W(+)], (F) Quintuple Dropout [2× AZ], (G) Sextuple Dropout [1× Pfizer], (H) Septuple Dropout [2x AZ] and (I) Octuple Dropout [2× AZ].

The cohort distribution antibody variant spectra are shown in Figure 2 as bee-swarm plots with the boxes showing upper and lower quartiles centred on the median and the whiskers indicating the presence of outliers at 1.5x the interquartile range, with the whisker bar falling on the highest/lowest non-outlier data point. The vaccine response is targeted at the Wuhan variant, in the centre of the plot. The pre-vaccine infection cohort, Figure 2A, shows median antibody levels for Wuhan and Spike protein variants, including the Omicron sub-variants. These median levels are increased in double and booster vaccination groups, Figure 2(B and C)The full spectrum of median values are reported in Table S3 along with the interquartile ranges. The interquartile range for Wuhan, natural infection cohort is low, with antibody levels varying from 0.2 – 1.8 mg/L, doubling in the double-vaccinated cohort to 0.6 – 3.6 mg/L and rising further in the boosted cohort 2 – 14 mg/L. The plots do not show the personalised detail of responses, particularly super- or low- responders; the lower outliers are clustered at the detection limit (200 ng/mL) and the upper outliers are grouped at the top of the figure.

**Figure 2.**
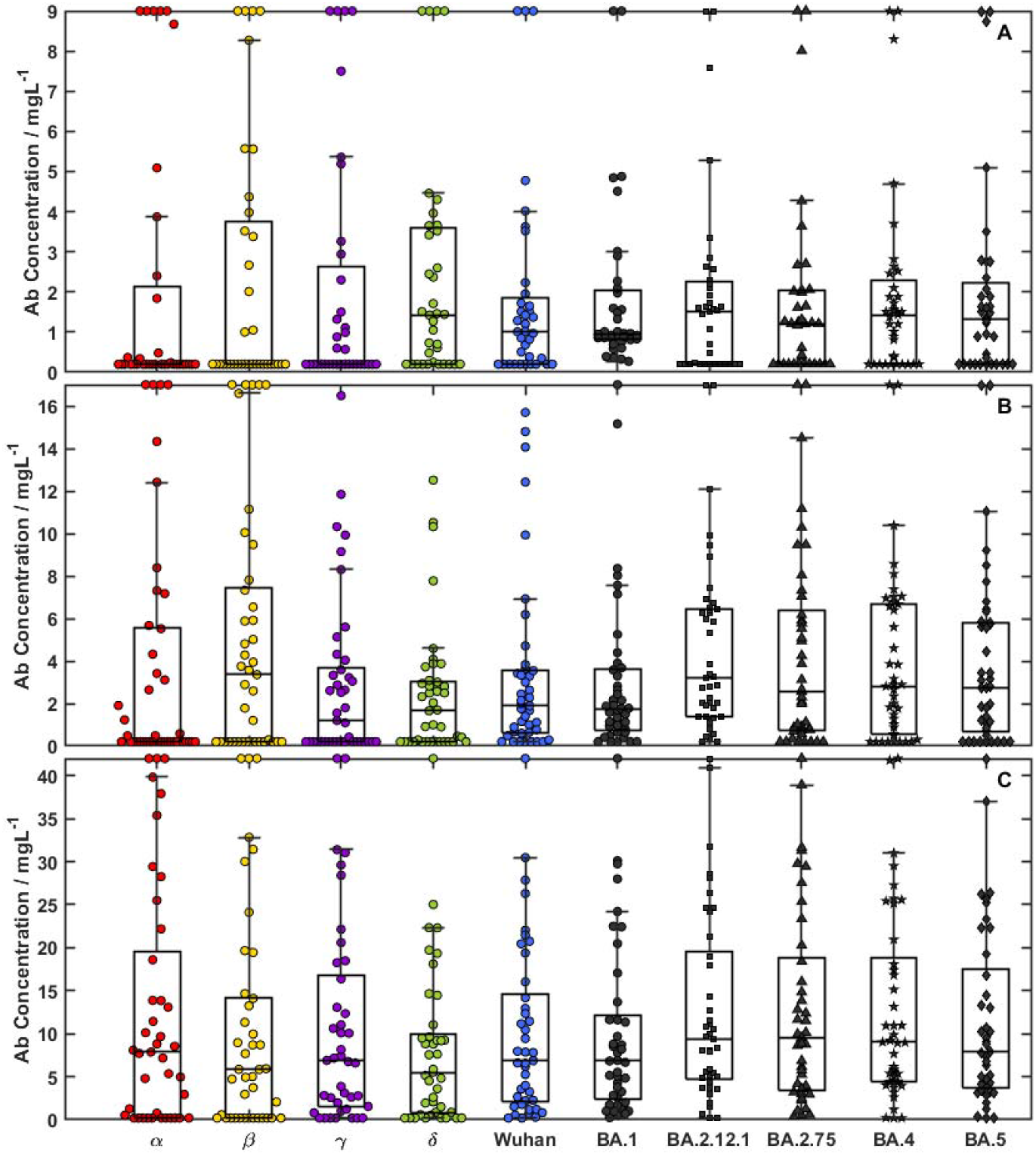
Bee swarm plots for 3 cohorts: (A) W(+); (B) double-vaccinated patients; and (C) triple-vaccinated patients. Outliers are shown at y-maximum and detailed in Table 1 outliers at 1.5× the interquartile range, with the whisker bar falling on the highest/lowest non-outlier data point.

Table 2 highlights patients responsible for larger responses, and their corresponding antibody concentrations for each Spike protein variant. Of these 17 super-responders, nine show a super-response (beyond the 95^th^ percentile of the population) to more than one variant and three show a full spectrum super-response.

**Table 2.** Outliers from Figure 2 detailed by patient, one per row.

| | $\alpha$ | $\beta$ | $\gamma$ | $\delta$ | $\mathbf{W}$ | $\mathbf{1}$ | $\mathbf{2.12.1}$ | $\mathbf{2.75}$ | $\mathbf{4}$ | $\mathbf{5}$ |
| --- | --- | --- | --- | --- | --- | --- | --- | --- | --- | --- |
| **W(+)** | 59.6 | 32.5 | 39.2 | 45.6 | 40.5 | 21.1 | 32.3 | 26.9 | 28.2 | 40.0 |
|  | 18.7 |  | 11.2 | 16.5 | 14.6 |  |  |  |  |  |
|  | 320 | 136.1 | 123.2 | 93.2 | 64.4 | 21.3 | 12.6 | 9.7 | 15.2 | 13.5 |
|  | 12.5 | 19.8 | 12.3 | 11.7 |  |  |  |  |  |  |
|  |  | 14.1 |  |  |  |  |  |  |  |  |
|  | 9.3 |  |  |  |  |  |  |  |  |  |
| **2x** | 28.1 | 19.7 |  |  |  |  |  |  |  |  |
| **Vax** | 52.8 | 50.9 |  |  |  |  |  |  |  |  |
|  |  |  |  |  |  |  | 27.5 | 26.9 | 26.2 | 25.1 |
|  | 74.2 | 40.2 |  |  |  |  |  |  |  |  |
|  |  | 29.5 |  |  |  |  |  |  |  |  |
|  | 18.9 |  |  |  |  | 20.7 | 29.4 | 27.3 | 29.8 | 27.8 |
|  |  | 18.6 |  |  |  |  |  |  |  |  |
| **3x** | 61.4 | 46.7 | 61.2 | 54.8 | 57.9 | 64.6 | 64.3 | 59.0 | 56.4 | 59.5 |
| **Vax** | 80.7 | 44.2 | 44.1 |  |  |  |  |  |  |  |
|  |  | 42.1 |  |  |  |  |  |  |  |  |
|  | 51.5 |  |  |  |  |  |  |  |  |  |

**Table 3.**
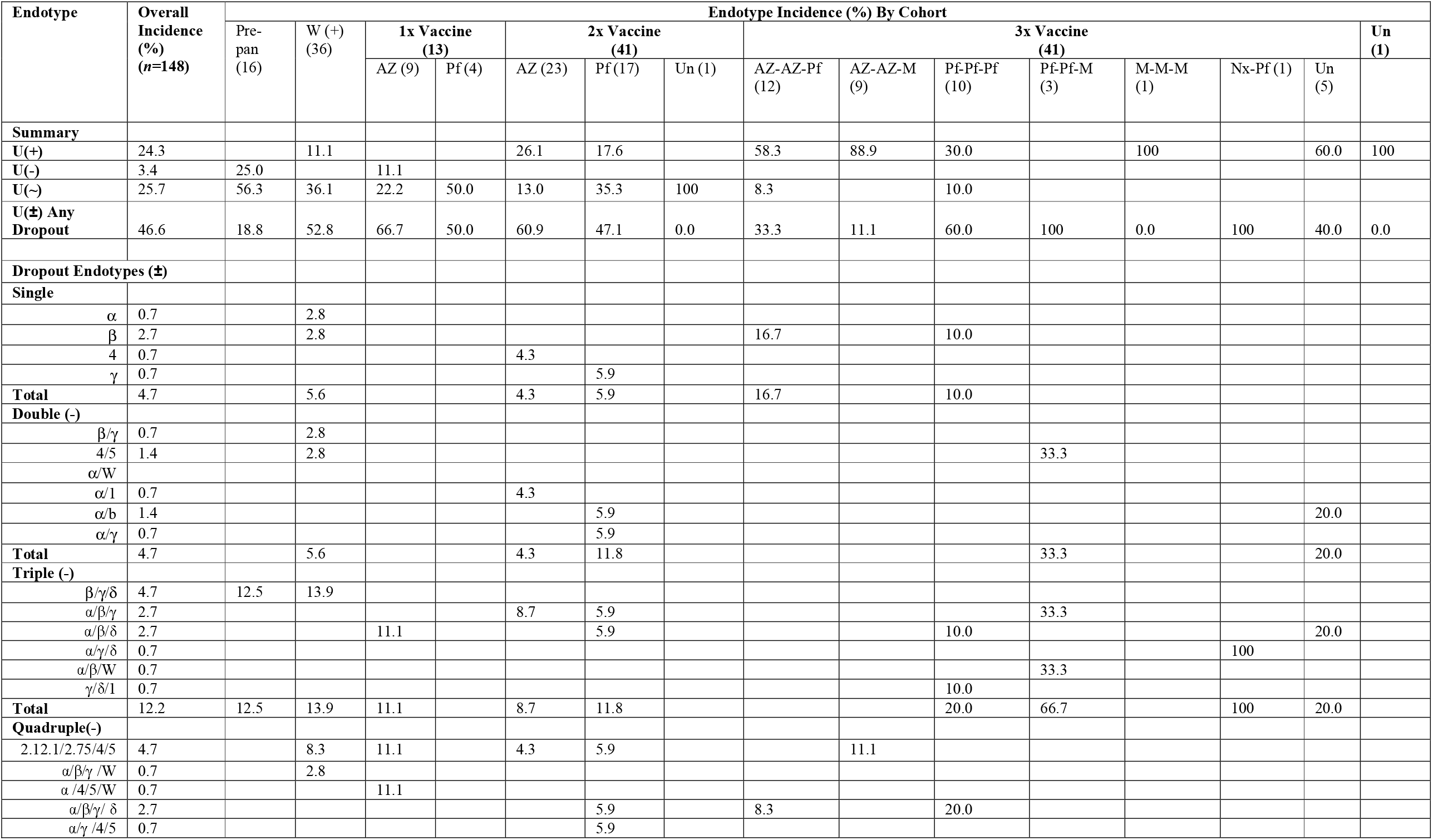
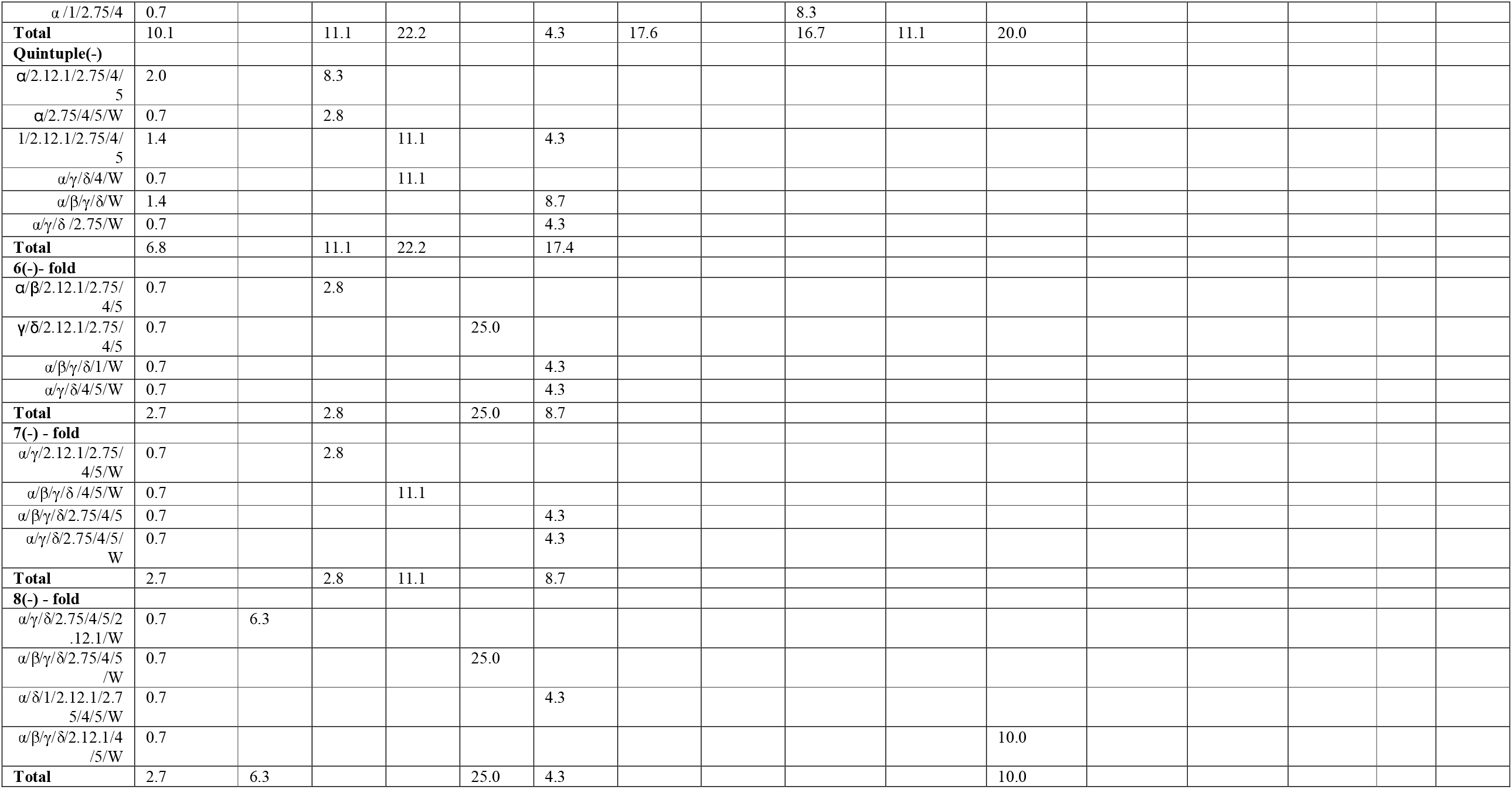
Incidence of endotype classifications for each of the patient cohorts. Un = undeclared data. Expressed as percentages wit number of samples in each cohort in shown in Table 1

## Discussion

A spectrum of antibody immunity across several of the SARS-CoV-2 variants has been investigated, with emphasis on the Omicron sub-variants that are currently or have most recently been in circulation (Figure S1). The antibody responses across cohorts were investigated; pre-vaccine, immunologically naïve patients; first vaccinations, and a fully mixed cohort of heterogeneous booster vaccines with immunity derived from vaccine alone, or in combination with previous infection. The pre-vaccine cohort shows antibody maturation in patients recovering from the infection with active clone selection during the serological response^16,17^. The extent of affinity maturation in the vaccination cohort is not well characterised but may be significantly smaller. The mixed booster cohort will however have antibodies from infection maturation as well as the original immunity imprint following vaccination. From September, 2022 there are four bivalent vaccines approved by the FDA^18^ providing protection against SARS-CoV-2: Pfizer-BioNTech COVID-19 Vaccine (Comirnaty®) – monovalent and bivalent (Original/Omicron BA.4/5); Moderna COVID-19 Vaccine (Spikevax®) - monovalent and bivalent (Original/Omicron BA.1); Johnson & Johnson’s Janssen COVID-19 Vaccine (Jcovden); and Novavax COVID-19 Vaccine (Nuvaxovid) based on adjuvanted Wuhan spike protein.

Each cohort shows a spectrum of immunity, Figure 2, with the median response concentration increasing from the W(+) cohort, through double vaccinated to boosted, with p-values from a Wilcoxon test in Table S6. The true insight however is in the individual personalised responses and distribution plots, with some evidence of bimodal distribution forming for some variants, although larger cohort sizes would be required to establish significance. The personalised spectral responses and the endotypes are useful for understanding the variation of immunity in the cohorts and ultimately the population.

The ideal antibody response produces a universal endotype U(+), with antibodies binding to a conserved endotype on all variants. The W(+), pre-vaccination cohort have antibodies matured against the epitope during an infection and show a U(+) incidence of 11% (95% CI 4% - 25%). Clone-selected antibodies are likely to be neutralising and bind to RBD or close to it to prevent entry into the cell and so are selected to the upper part of the S protein. Of the 53% (95% CI 37% - 68%) of patients showing drop-out endotypes U(±), some showed gaps in immunity for the early Alpha and Beta variants but more significantly towards the Omicron variants. The U(±) immunity imprints acquired during this initial infection may lead to B cell memory that does not respond to subsequent vaccination or infection from new variants. The ‘one-and-done’ idea of immunity acquired from one infection will protect against future infections may be true for the 11% U(+) endotypes but is more likely to be false for the majority of the population.

The double-vaccination cohort U(+) incidence was 22% (95% CI 12% - 37%) and 54% (95% CI 39% - 68%) in the triple vaccinated cohort. These antibodies are not matured against infection clearance efficacy and could be raised to any part of the spike protein and with lower affinity. The U(±) immunity heterogeneity was greatest for the double-vaccinated AZ cohort, and was subsequently reduced with the booster, implying the booster broke the immunity imprint provided by the initial vaccination. This provides hope that a smart-boosting strategy could improve patient immunity, particularly towards future variants. It is worth note that the triple-vaccinated cohort could also have a mixture of infection-matured antibodies that have been enhanced by the vaccines.

The mutations to the Wuhan spike protein are shown in Figure 3(A) and the same protein extended with the Omicron variants in Figure 3(B). The regions showing the most mutations are the targets of RBD neutralising antibodies and ACE2 receptor binding, but the hinge remains untouched. The S2 region also shows a far lower mutation density.

**Figure 3.**
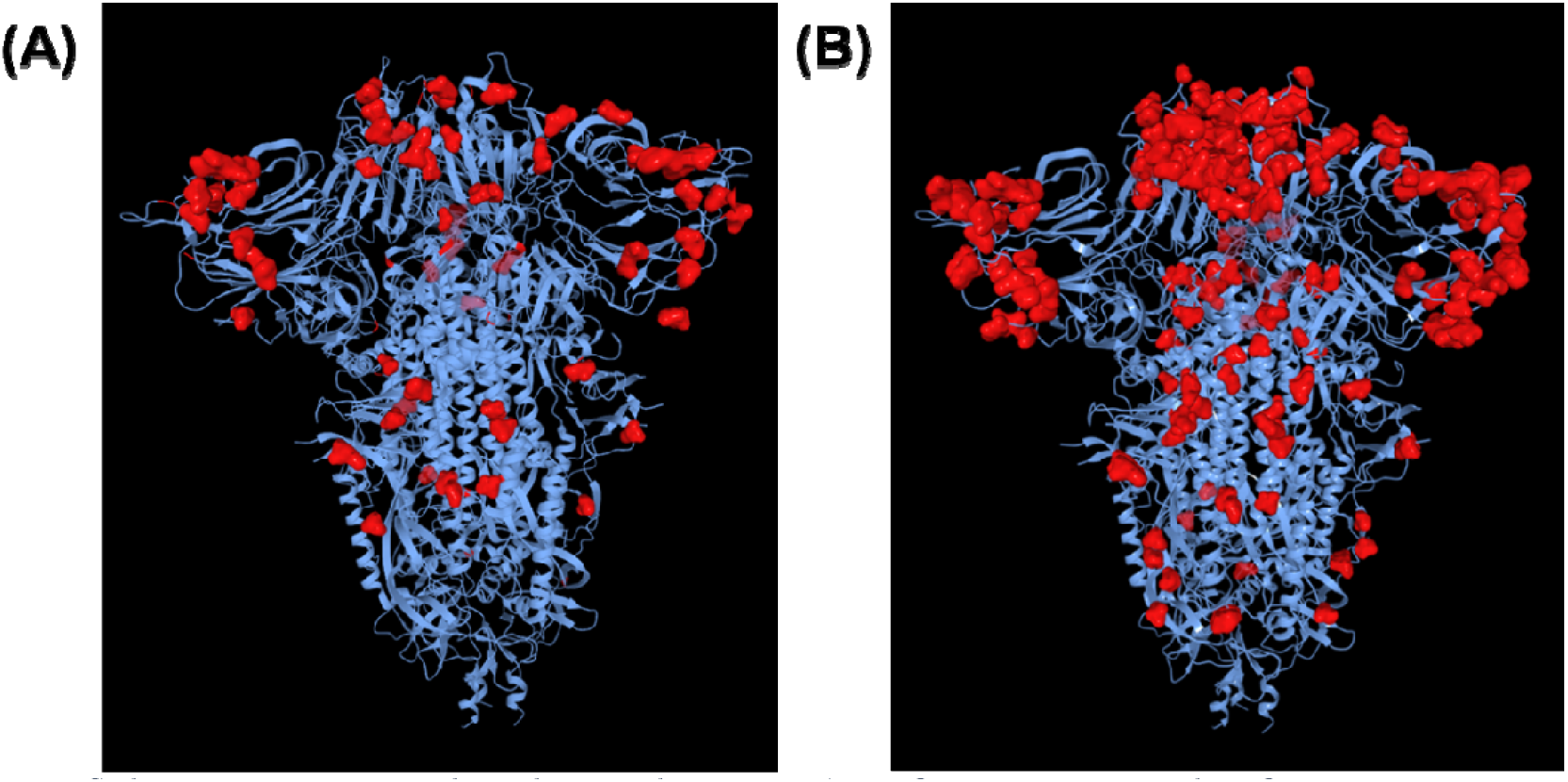
Spike Protein mutations on the Wuhan initial structure: (A) pre-Omicron variants; and (B) Omicron variants (without BA 2.75)

The antibody immunity conferred on each variant can be measured directly for new variants as they arise, but the response to BA.1 appears to predict the response to the other Omicron sub-variants with relative accuracy; no single sub-variant showed a singular drop-out, also showing high correlation, as shown in Table S4. Whilst these predictions do not capture the individual variant spectrums, it can provide insight into population immunity on a national, or potentially global scale.

The duration of the vaccines efficacy against infection and serious disease has been reviewed systematically^24^, but does not quantify the variation seen in immunity endotypes for individuals within cohorts and their risk against any new evolution of the virus. Some in the U(+) cohort may continue to show protection from infection if antibody levels are maintained, and if the antibodies produced are of high affinity. However, those with a full vaccination history such as the triple-Pfizer octuple drop-out α/β/γ/δ/2.12.1/4/5/W may have little or no protection to the currently circulating variants, increasing risk to recurrent infections, potentially from failure to break to imprinted immunity endotype.

Elimination of an infection requires both antibody and T cells to complete clearance of the virus from the localised infection site, but systemic clearance requires a sterilising serum^19^, where antibodies alone provide elimination of the virus. Poor quality antibodies permeating throughout the body, possibly associated with a U(±) endotype, may allow persistence of the SARS-CoV-2 virus in micro-colonies. Viral persistence has been linked to the pathophysiology of long covid^20^ which may explain some of the collection of symptoms observed in affected individuals^21^. The U(±) incidence in the triple vaccination cohort is 44% (95% CI 30% - 57%). This can allow for viral persistence in a significant proportion of the population, which is currently recorded in the UK as much as one-in-five infections^22^. The consequences for UK workforce^23^ especially in healthcare and teaching may lead to a significant burden. Long Covid patients early in the development of symptoms may benefit from a smart boost of immunotherapy or a different vaccine, as seen in the triple booster cohort, to compensate from the immunity endotype. The drop-out endotype may provide a pathophysiology for long Covid.

## Conclusions

The antibody immunity spectrum has shown a prevalence of the U(+) universal endotype to a common epitope on all variants and points to the prospect for a universal vaccine or immunotherapy against SARS-CoV-2. Less encouraging is the U(±) incidence of 41% (95% CI 28% - 57%), which suggests that nearly two-thirds of the population may have an endotype that will not confer protection or worse, allow persistence of the virus with a non-sterilising serum. A smart boosting campaign with different types of vaccines^10,11^ may be required to break the immunity imprint by choosing heterogeneous vaccine sequences, along with the utilisation of new bivalent or whole-virus vaccines.

## Data Availability

All data produced in the present study are available upon reasonable request to the authors

## Acknowledgements

The authors would like to thank Dr Jonathan Snicker for their guidance on the potential strategic and policy implications of the scientific findings.

## Supplementary Data

**Table S1.**
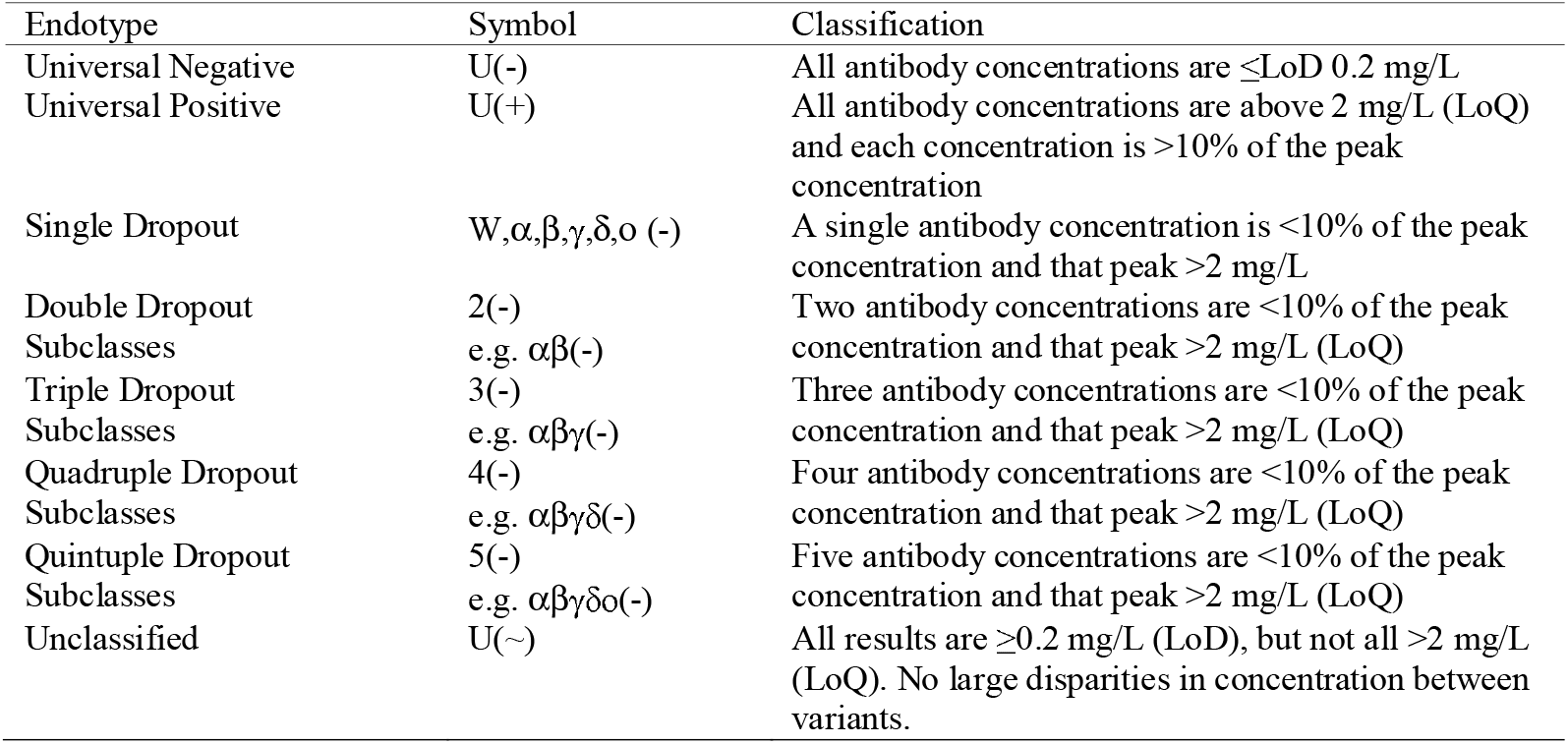
Endotype classification and symbols used to represent them.

**Table S2.** Spike protein details, full mutation lists can be found on the relevant product datasheets.

| Protein | Supplier | Cat # | Lot # | Type | Cell Line | Tag | AA Length |
| --- | --- | --- | --- | --- | --- | --- | --- |
| Wuhan | Sinobiological | 40589-V08B1 | MF16JU1612 | Monomer | Baculovirus-Insect | C-term polyhistidine | 1209 |
| Alpha | Sinobiological | 40589-V08B6 | LC15MC0903 | Monomer | Baculovirus-Insect | C-term polyhistidine | 1206 |
| Beta | Sinobiological | 40589-V08B7 | LC15MY2607 | Monomer | Baculovirus-Insect | C-term polyhistidine | 1206 |
| Gamma | Sinobiological | 40589-V08B8 | LC15JU0209 | Monomer | Baculovirus-Insect | C-term polyhistidine | 1209 |
| Delta | Sinobiological | 40589-V08B16 | LC15OC2012 | Monomer | Baculovirus-Insect | C-term polyhistidine | 1207 |
| Omicron | Sinobiological | 40589-V08B33 | MF15DE2961 | Monomer | Baculovirus-Insect | C-term polyhistidine | 1206 |
| BA.1 | Sinobiological | 40589-V08H26 | LC16JA1515 | Trimer | HEK293 | C-term polyhistidine | 1228 |
| BA.2.12.1 | Sinobiological | 40589-V08H34 | MB16JU1681 | Trimer | HEK293 | C-term polyhistidine | 1228 |
| BA.2.75 | Acro Biosystems | SPN-C522f | 6764A-227DF1-146 | Trimer | HEK293 | C-term polyhistidine | 1197 |
| BA.4 | Sinobiological | 40589-V08H32 | MB16JU1382 | Trimer | HEK293 | C-term polyhistidine | 1226 |
| BA.5 | Sinobiological | 40589-V08H33 | MB16JU1383 | Trimer | HEK293 | C-term polyhistidine | 1226 |

**Table S3.** Table of medians and quartiles, broken down by SARS-CoV-2 exposure cohort.

| Pre-Pandemic | $\alpha$ | $\beta$ | $\gamma$ | $\delta$ | Wuhan | BA.1 | BA.2.12.1 | BA.2.75 | BA.4 | BA.5 |
| --- | --- | --- | --- | --- | --- | --- | --- | --- | --- | --- |
| Lower Quartile | 0.2 | 0.2 | 0.2 | 0.2 | 0.2 | 0.2 | 0.2 | 0.2 | 0.2 | 0.2 |
| Median | 0.2 | 0.2 | 0.2 | 0.2 | 0.2 | 0.4 | 0.2 | 0.2 | 0.3 | 0.3 |
| Upper Quartile | 0.6 | 0.2 | 0.2 | 0.2 | 0.2 | 0.7 | 1.0 | 0.7 | 0.9 | 0.9 |
| Wuhan (+) |  |  |  |  |  |  |  |  |  |  |
| Lower Quartile | 0.2 | 0.2 | 0.2 | 0.2 | 0.2 | 0.8 | 0.2 | 0.2 | 0.2 | 0.2 |
| Median | 0.2 | 0.2 | 0.2 | 1.4 | 1.0 | 1.0 | 1.5 | 1.2 | 1.4 | 1.3 |
| Upper Quartile | 2.1 | 3.8 | 2.6 | 3.6 | 1.8 | 2.0 | 2.2 | 2.0 | 2.3 | 2.2 |
| Double Vaccinated |  |  |  |  |  |  |  |  |  |  |
| Lower Quartile | 0.2 | 0.2 | 0.2 | 0.2 | 0.6 | 0.8 | 1.4 | 0.7 | 0.5 | 0.7 |
| Median | 0.2 | 3.4 | 1.2 | 1.7 | 1.9 | 1.8 | 3.2 | 2.6 | 2.8 | 2.8 |
| Upper Quartile | 5.6 | 7.5 | 3.7 | 3.0 | 3.6 | 3.6 | 6.5 | 6.4 | 6.7 | 5.8 |
| Triple Vaccinated |  |  |  |  |  |  |  |  |  |  |
| Lower Quartile | 0.2 | 0.2 | 1.5 | 0.8 | 2.1 | 2.4 | 4.8 | 3.5 | 4.4 | 3.7 |
| Median | 7.9 | 5.8 | 6.9 | 5.5 | 6.9 | 6.8 | 9.4 | 9.6 | 9.0 | 7.9 |
| Upper Quartile | 19.5 | 14.2 | 16.9 | 10.0 | 14.6 | 12.1 | 19.5 | 18.9 | 18.8 | 17.4 |

**Table S4.** Table of Line-of-best fit gradients, column = row × gradient +intercept. The intercept is the Limit of Detection.

| Wuhan (+) | $\alpha$ | $\beta$ | $\gamma$ | $\delta$ | Wuhan | BA.1 | BA.2.12.1 | BA.2.75 | BA.4 | BA.5 |
| --- | --- | --- | --- | --- | --- | --- | --- | --- | --- | --- |
| $\alpha$ | 1.00 | 0.42 | 0.39 | 0.30 | 0.21 | 0.07 | 0.05 | 0.04 | 0.06 | 0.05 |
| $\beta$ | 2.30 | 1.00 | 0.91 | 0.71 | 0.50 | 0.17 | 0.12 | 0.10 | 0.13 | 0.12 |
| $\gamma$ | 2.51 | 1.08 | 1.00 | 0.79 | 0.56 | 0.19 | 0.15 | 0.12 | 0.16 | 0.15 |
| $\delta$ | 3.04 | 1.31 | 1.23 | 1.00 | 0.73 | 0.26 | 0.23 | 0.19 | 0.24 | 0.23 |
| Wuhan | 3.98 | 1.72 | 1.64 | 1.35 | 1.00 | 0.37 | 0.35 | 0.28 | 0.35 | 0.35 |
| BA.1 | 9.09 | 4.02 | 3.89 | 3.32 | 2.51 | 1.00 | 1.05 | 0.86 | 1.01 | 1.04 |
| BA.2.12.1 | 4.52 | 2.06 | 2.16 | 2.09 | 1.70 | 0.75 | 1.00 | 0.83 | 0.91 | 0.98 |
| BA.2.75 | 5.07 | 2.32 | 2.47 | 2.42 | 1.99 | 0.88 | 1.19 | 1.00 | 1.09 | 1.17 |
| BA.4 | 5.93 | 2.62 | 2.70 | 2.51 | 2.01 | 0.85 | 1.07 | 0.89 | 1.00 | 1.06 |
| BA.5 | 4.90 | 2.19 | 2.30 | 2.20 | 1.78 | 0.77 | 1.02 | 0.85 | 0.94 | 1.00 |
| **Double Vaccinated** |  |  |  |  |  |  |  |  |  |  |
| $\alpha$ | 1.00 | 0.64 | 0.14 | 0.14 | 0.24 | 0.14 | 0.11 | 0.13 | 0.11 | 0.11 |
| $\beta$ | 1.11 | 1.00 | 0.18 | 0.13 | 0.25 | 0.13 | 0.06 | 0.08 | 0.06 | 0.07 |
| $\gamma$ | 2.03 | 1.42 | 1.00 | 0.67 | 0.76 | 0.79 | 0.90 | 0.90 | 0.94 | 0.89 |
| $\delta$ | 3.30 | 1.83 | 1.12 | 1.00 | 1.26 | 1.15 | 1.38 | 1.40 | 1.39 | 1.32 |
| Wuhan | 2.87 | 1.74 | 0.65 | 0.64 | 1.00 | 0.81 | 1.00 | 1.07 | 1.01 | 0.97 |
| BA.1 | 1.72 | 0.94 | 0.71 | 0.61 | 0.85 | 1.00 | 1.38 | 1.38 | 1.35 | 1.30 |
| BA.2.12.1 | 0.61 | 0.20 | 0.36 | 0.33 | 0.47 | 0.62 | 1.00 | 1.00 | 0.98 | 0.94 |
| BA.2.75 | 0.72 | 0.26 | 0.34 | 0.32 | 0.48 | 0.59 | 0.95 | 1.00 | 0.95 | 0.92 |
| BA.4 | 0.62 | 0.19 | 0.37 | 0.33 | 0.47 | 0.60 | 0.98 | 1.00 | 1.00 | 0.95 |
| BA.5 | 0.68 | 0.25 | 0.39 | 0.35 | 0.50 | 0.64 | 1.03 | 1.06 | 1.04 | 1.00 |
| **Triple Vaccinated** |  |  |  |  |  |  |  |  |  |  |
| $\alpha$ | 1.00 | 0.57 | 0.67 | 0.45 | 0.50 | 0.49 | 0.48 | 0.48 | 0.46 | 0.45 |
| $\beta$ | 1.11 | 1.00 | 0.79 | 0.56 | 0.61 | 0.59 | 0.56 | 0.56 | 0.53 | 0.52 |
| $\gamma$ | 1.27 | 0.77 | 1.00 | 0.73 | 0.79 | 0.83 | 0.81 | 0.80 | 0.76 | 0.75 |
| $\delta$ | 1.46 | 0.91 | 1.24 | 1.00 | 1.08 | 1.16 | 1.15 | 1.12 | 1.06 | 1.06 |
| Wuhan | 1.32 | 0.82 | 1.10 | 0.89 | 1.00 | 1.04 | 1.06 | 1.05 | 1.00 | 0.98 |
| BA.1 | 1.14 | 0.69 | 1.00 | 0.83 | 0.91 | 1.00 | 1.02 | 0.99 | 0.94 | 0.94 |
| BA.2.12.1 | 1.00 | 0.59 | 0.89 | 0.75 | 0.84 | 0.92 | 1.00 | 0.97 | 0.94 | 0.92 |
| BA.2.75 | 1.02 | 0.61 | 0.90 | 0.75 | 0.85 | 0.92 | 1.00 | 1.00 | 0.96 | 0.92 |
| BA.4 | 1.06 | 0.62 | 0.92 | 0.76 | 0.86 | 0.94 | 1.04 | 1.03 | 1.00 | 0.96 |
| BA.5 | 1.09 | 0.65 | 0.97 | 0.81 | 0.91 | 1.00 | 1.08 | 1.06 | 1.03 | 1.00 |

**Table S5.** Table of R^2^ values.

| Wuhan (+) | $\alpha$ | $\beta$ | $\gamma$ | $\delta$ | Wuhan | BA.1 | BA.2.12.1 | BA.2.75 | BA.4 | BA.5 |
| --- | --- | --- | --- | --- | --- | --- | --- | --- | --- | --- |
| $\alpha$ | 1.00 | 0.99 | 0.99 | 0.96 | 0.92 | 0.80 | 0.47 | 0.44 | 0.57 | 0.50 |
| $\beta$ | 0.99 | 1.00 | 0.99 | 0.96 | 0.92 | 0.83 | 0.50 | 0.47 | 0.59 | 0.52 |
| $\gamma$ | 0.99 | 0.99 | 1.00 | 0.98 | 0.96 | 0.87 | 0.57 | 0.55 | 0.66 | 0.60 |
| $\delta$ | 0.96 | 0.96 | 0.98 | 1.00 | 0.99 | 0.93 | 0.69 | 0.67 | 0.77 | 0.71 |
| Wuhan | 0.92 | 0.92 | 0.96 | 0.99 | 1.00 | 0.96 | 0.77 | 0.75 | 0.84 | 0.79 |
| BA.1 | 0.80 | 0.83 | 0.87 | 0.93 | 0.96 | 1.00 | 0.89 | 0.87 | 0.92 | 0.89 |
| BA.2.12.1 | 0.47 | 0.50 | 0.57 | 0.69 | 0.77 | 0.89 | 1.00 | 1.00 | 0.99 | 1.00 |
| BA.2.75 | 0.44 | 0.47 | 0.55 | 0.67 | 0.75 | 0.87 | 1.00 | 1.00 | 0.99 | 0.99 |
| BA.4 | 0.57 | 0.59 | 0.66 | 0.77 | 0.84 | 0.92 | 0.99 | 0.99 | 1.00 | 1.00 |
| BA.5 | 0.50 | 0.52 | 0.60 | 0.71 | 0.79 | 0.89 | 1.00 | 0.99 | 1.00 | 1.00 |
| **Double Vaccinated** |  |  |  |  |  |  |  |  |  |  |
| $\alpha$ | 1.00 | 0.84 | 0.54 | 0.68 | 0.82 | 0.48 | 0.26 | 0.31 | 0.26 | 0.27 |
| $\beta$ | 0.84 | 1.00 | 0.50 | 0.50 | 0.66 | 0.35 | 0.11 | 0.15 | 0.11 | 0.13 |
| $\gamma$ | 0.54 | 0.50 | 1.00 | 0.86 | 0.70 | 0.75 | 0.57 | 0.56 | 0.59 | 0.59 |
| $\delta$ | 0.68 | 0.50 | 0.86 | 1.00 | 0.90 | 0.84 | 0.67 | 0.67 | 0.68 | 0.68 |
| Wuhan | 0.82 | 0.66 | 0.70 | 0.90 | 1.00 | 0.83 | 0.68 | 0.72 | 0.69 | 0.69 |
| BA.1 | 0.48 | 0.35 | 0.75 | 0.84 | 0.83 | 1.00 | 0.92 | 0.90 | 0.90 | 0.91 |
| BA.2.12.1 | 0.26 | 0.11 | 0.57 | 0.67 | 0.68 | 0.92 | 1.00 | 0.97 | 0.98 | 0.98 |
| BA.2.75 | 0.31 | 0.15 | 0.56 | 0.67 | 0.72 | 0.90 | 0.97 | 1.00 | 0.98 | 0.98 |
| BA.4 | 0.26 | 0.11 | 0.59 | 0.68 | 0.69 | 0.90 | 0.98 | 0.98 | 1.00 | 1.00 |
| BA.5 | 0.27 | 0.13 | 0.59 | 0.68 | 0.69 | 0.91 | 0.98 | 0.98 | 1.00 | 1.00 |
| **Triple Vaccinated** |  |  |  |  |  |  |  |  |  |  |
| $\alpha$ | 1.00 | 0.79 | 0.92 | 0.81 | 0.81 | 0.75 | 0.69 | 0.70 | 0.70 | 0.70 |
| $\beta$ | 0.79 | 1.00 | 0.78 | 0.71 | 0.71 | 0.64 | 0.57 | 0.58 | 0.57 | 0.58 |
| $\gamma$ | 0.92 | 0.78 | 1.00 | 0.96 | 0.94 | 0.91 | 0.85 | 0.85 | 0.84 | 0.86 |
| $\delta$ | 0.81 | 0.71 | 0.96 | 1.00 | 0.98 | 0.98 | 0.93 | 0.92 | 0.90 | 0.93 |
| Wuhan | 0.81 | 0.71 | 0.94 | 0.98 | 1.00 | 0.97 | 0.94 | 0.95 | 0.93 | 0.94 |
| BA.1 | 0.75 | 0.64 | 0.91 | 0.98 | 0.97 | 1.00 | 0.97 | 0.96 | 0.94 | 0.97 |
| BA.2.12.1 | 0.69 | 0.57 | 0.85 | 0.93 | 0.94 | 0.97 | 1.00 | 0.98 | 0.99 | 1.00 |
| BA.2.75 | 0.70 | 0.58 | 0.85 | 0.92 | 0.95 | 0.96 | 0.98 | 1.00 | 0.99 | 0.99 |
| BA.4 | 0.70 | 0.57 | 0.84 | 0.90 | 0.93 | 0.94 | 0.99 | 0.99 | 1.00 | 0.99 |
| BA.5 | 0.70 | 0.58 | 0.86 | 0.93 | 0.94 | 0.97 | 1.00 | 0.99 | 0.99 | 1.00 |

**Table S6.**
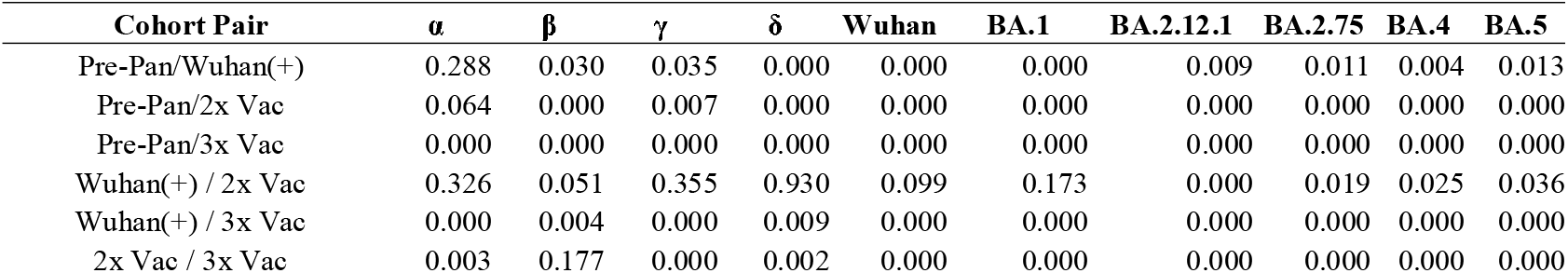
Table of p-values from a Wilcoxon test comparing the medians of each cohort on a by-channel basis.

| Cohort Pair | $\alpha$ | $\beta$ | $\gamma$ | $\delta$ | Wuhan | BA.1 | BA.2.12.1 | BA.2.75 | BA.4 | BA.5 |
| --- | --- | --- | --- | --- | --- | --- | --- | --- | --- | --- |
| Pre-Pan/Wuhan(+) | 0.288 | 0.030 | 0.035 | 0.000 | 0.000 | 0.000 | 0.009 | 0.011 | 0.004 | 0.013 |
| Pre-Pan/2x Vac | 0.064 | 0.000 | 0.007 | 0.000 | 0.000 | 0.000 | 0.000 | 0.000 | 0.000 | 0.000 |
| Pre-Pan/3x Vac | 0.000 | 0.000 | 0.000 | 0.000 | 0.000 | 0.000 | 0.000 | 0.000 | 0.000 | 0.000 |
| Wuhan(+)/2x Vac | 0.326 | 0.051 | 0.355 | 0.930 | 0.099 | 0.173 | 0.000 | 0.019 | 0.025 | 0.036 |
| Wuhan(+)/3x Vac | 0.000 | 0.004 | 0.000 | 0.009 | 0.000 | 0.000 | 0.000 | 0.000 | 0.000 | 0.000 |
| 2x Vac/3x Vac | 0.003 | 0.177 | 0.000 | 0.002 | 0.000 | 0.000 | 0.000 | 0.000 | 0.000 | 0.000 |

**Figure S1.**
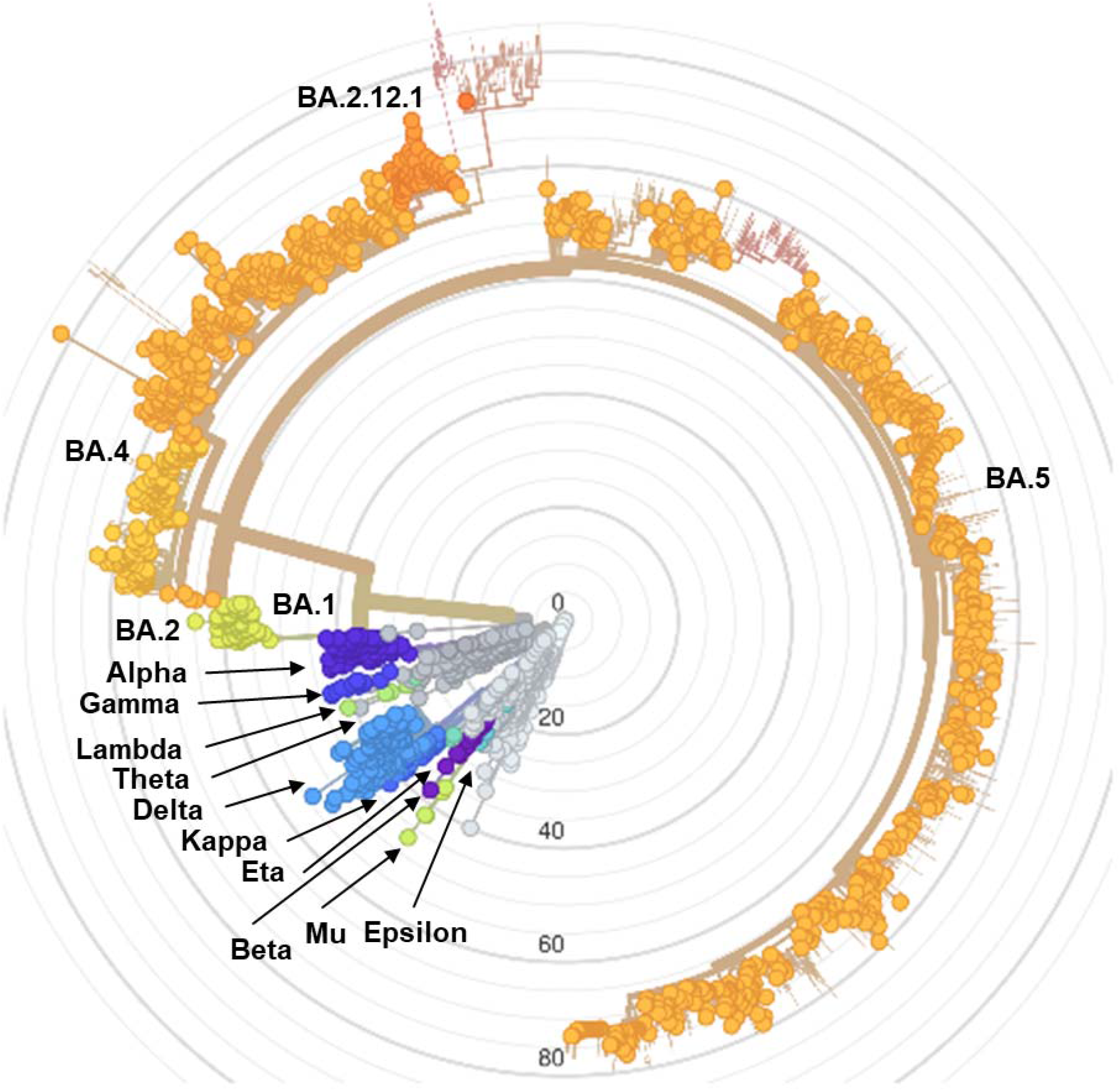
SARS-CoV-2 genetic divergence tree diagram. Radial axis is ‘number of mutations from first SARS-CoV-2 sequence’. Created using Nextstrain.^25^

